# “It Just Puts You Out!” ‘Tranq Sleep,’ Sensory Discernment, and the Experience of Consuming Xylazine-Adulterated Fentanyl in Rural North Carolina

**DOI:** 10.64898/2026.10.30.26366218

**Authors:** Claudia Squire, Aminah Isiaq, Nicholas Peiper, Michelle Mathis, Jon E Zibbell

## Abstract

**Background:** Xylazine, a veterinary sedative not approved for human use, has emerged as a consequential adulterant in the U.S. illicit fentanyl supply. Although a growing body of qualitative research has documented the experiences of fentanyl consumers exposed to ‘tranq’ in large Northeastern cities, little is known about how xylazine is experienced in rural or Southern regions where the evolution of the illicit drug supply often lags behind its urban equivalents. This study examines how fentanyl consumers in west-central North Carolina detect, comprehend and respond to the widespread adulteration of fentanyl markets with xylazine.

**Methods:** Between September and December of 2025, a seasoned interviewer conducted 25 in-depth, semi-structured qualitative interviews with fentanyl consumers recruited from both fixed and mobile syringe service programs in west-central North Carolina. Interviews explored sensory discernment strategies, pharmacologic effects, and behavioral responses to the presence of xylazine. Data were analyzed using a combined deductive-inductive thematic approach in QSR NVivo 14.0.

**Results:** Participants described a distinctive repertoire of sensory strategies for identifying xylazine in their fentanyl supply, including the appearance of drug residue on foil, bitterness and chemical odor when smoked, and altered viscosity when injected. A minority described an inability to detect xylazine through sensory cues. Participants consistently distinguished a profound involuntary sedation they called “tranq sleep” from the familiar opioid nod, characterizing it by nearly instantaneous loss of consciousness with severe withdrawal upon awakening. While nearly all participants expressed strong aversion to xylazine, a meaningful minority valued its amplifying effects to ‘improve’ a diminishing fentanyl high.

**Conclusions:** This study offers among the first qualitative description of xylazine exposure in a rural fentanyl market. Findings extend the xylazine literature geographically and pharmacologically and underscore the urgency of harm reduction responses tailored to the distinctive harms of xylazine. In particular, adverse health outcomes that accumulate during episodes of protracted and unarousable sedation, including physical injury, victimization, and the severe withdrawal syndrome that follows upon awakening.

**Highlights:** xylazine; tranq sleep, fentanyl; qualitative methods; harm reduction; rural drug markets; drug discernment

## Introduction

The United States opioid epidemic has advanced through several distinct but intersecting waves each defined by a shift in the composition and lethality of the illicit opioid supply. The most recent transformation involves not only the market displacement of heroin by illicitly manufactured fentanyls (hereafter fentanyl) but the emergence of novel synthetic depressants that are reshaping the experience of fentanyl consumption (Sisco and Appley, 2023). Among the most current and consequential is xylazine, an alpha-2 adrenergic agonist not approved for human use and colloquially referred to as *tranq* for the potent sedation it delivers (Ball et al., 2022). First documented during the current opioid epidemic in Philadelphia as an adulterant of fentanyl, xylazine has since spread into illicit markets throughout the continental U.S. and is now recognized as a public health emergency (Kariisa et al., 2023).

Xylazine’s emergence as a public health threat is inseparable from the fentanyl supply that carries it. In one of the earliest national syntheses, Friedman and colleagues (2022) documented a pattern of increasing co-occurrence of xylazine in fentanyl overdose deaths across multiple U.S. jurisdictions, establishing its epidemiologic footprint before the clinical consequences of human exposure (e.g., necrotic wounds) were well understood. As the xylazine-adulterated fentanyl supply matured and spread geographically, qualitative and mixed-methods research began to characterize how consumers perceived and were responding to its unwanted presence. The portrait that emerged was consistent and striking in its uniformity: fentanyl consumers do not want tranq.

Reed and colleagues (2022) found a collective disdain among fentanyl consumers in Philadelphia who viewed xylazine as a deeply undesirable adulterant, one they detested for its effects, feared for the safety vulnerabilities it created, and actively sought to avoid whenever possible. This aversion is not a regional phenomenon specific to Philadelphia but a consistent feature of the xylazine experience wherever it has been closely examined. A community drug-checking study in Rhode Island found fentanyl consumers universally attempting to avoid xylazine, having learned to rely on its extreme sedating effects as an *ex post facto* signal of a presence they could not have detected prior to consumption (Ujeneza et al., 2025).

Despite the recent growth in qualitative xylazine research, a significant geographic gap persists in our understanding of the consumer experience outside of Northeastern and MidAtlantic cities. Nearly all research on xylazine has been conducted in Philadelphia, Baltimore, Providence or comparable urban markets where xylazine penetrated earliest and with the greatest intensity, leaving rural and Southern drug markets largely unexamined. The present study addresses this gap by examining how fentanyl consumers in the Appalachian Foothills of west-central North Carolina are detecting and responding to xylazine in rural settings where formal drug-checking infrastructure remains largely unavailable to consumers.

## Methods

### Study Setting and Recruitment

This study was conducted in west-central North Carolina (NC), a predominantly rural region in the state’s Appalachian foothills. Recruitment was conducted in close partnership with Olive Branch Ministry (OBM), a faith-based, syringe services program. OBM provides harm reduction services to people who inject drugs (PWID) via two storefront locations and a mobile outreach program for the more geographically isolated (hinterland) communities. Study participants were recruited from locations in OBM’s service area and included fixed and mobile program sites, home locations, and wooded encampments.

Individuals were invited to complete a one-time, 60-minute qualitative interview to share their knowledge of and experience consuming xylazine. Participants received $30 cash stipend as honoraria for their time and knowledge sharing. Eligibility criteria involved being at least 18 years of age, residing in North Carolina, ever consuming xylazine, using fentanyl in the past 48 hours, proficiency in spoken English, and able to provide informed consent. OBM provided a private setting for interview activities not conducted *in situ*.

### Data Collection

Between September and December 2025, the Principal Investigator conducted and digitally audio recorded all interviews. The semi-structured interview guide was developed from his protracted ethnographic fieldwork in west-central North Carolina and covered four substantive domains: sensory discernment strategies for detecting xylazine in the fentanyl supply; the pharmacologic effects of xylazine as experienced by consumers; comparative assessments of xylazine relative to heroin and fentanyl; and partialities for xylazine-adulterated fentanyl. The depth and specificity of the guide’s domains reflects years of direct observation and relationship-building in the PWID community. All participants provided oral informed consent prior to interview. Audio recordings were professionally transcribed verbatim, de-identified, and uploaded to a secure database for qualitative analysis. The study received full board review and approval from the Institutional Review Board at RTI International.

### Data Analysis

Participants were anonymized and tracked in a database alongside demographic information and drug sample results where applicable. De-identified transcripts were uploaded into QSR NVivo 14 for coding and analysis. An initial codebook of deductive codes was derived from topics in the interview guide, itself developed from ethno-epidemiological fieldwork conducted in the study locality over many years (Erickson et al., 2026; Peiper et al., 2025; Zibbell et al., 2024; Duhart Clarke et al., 2022; Zibbell et al, 2021; Peiper et al., 2019). Two team members independently coded transcripts until intercoder reliability reached Cohen’s Kappa ≥ 0.80 across all codes; remaining transcripts were then divided between coders. Throughout the coding process the codebook was refined and inductive codes were added to capture themes emerging from the data that were not anticipated by the guide. Thematic analysis was then employed to develop narrative summaries of salient findings and construct quote tables around each theme. During analysis, participant characteristics—including recent illicit stimulant use and primary route of fentanyl consumption—were added to the participant database to support within-sample comparisons. The final analytical sample comprised 25 participants.

## RESULTS

Participants ranged widely in age (28 - 67 years; mean 39) and were predominantly non-Hispanic White, with everyone residing in either Gaston (n=19) or neighboring Burke County (n=6). Concurrent patterns of illicit opioid and stimulant consumption reflected rural North Carolina drug markets as previously documented by the study team (Peiper et al., 2025). Most participants report smoking as their primary route of fentanyl consumption but all participants continued to inject drugs. Three primary themes related to xylazine exposure emerged across interviews: (1) xylazine’s physical effect as a distinct form of acute unconscious sedation; (2) the capacity to recognize xylazine in fentanyl by sensory discernment strategies; and (3) prevailing attitudes and preferences toward xylazine.

### Tranq Sleep

A consistent and prominent finding was xylazine’s distinct form of tranquilizing sedation when smoked or injected. Participants described this sedation as a near-instantaneous collapse into an anesthetized state from which they could not be easily roused. Participants uniformly distinguished this acute sedation from the familiar opioid “nod” they sought from and experienced with heroin. We label this phenomenon “*tranq sleep*” to capture the abrupt, involuntary, and incapacitating nature of the sedation explained by participants. Unlike the gradual and cognizant drowsiness of an opioid ’nod,’ *tranq sleep* was described as a sudden, binary transition from full wakefulness to deep sleep with no warning phase and no capacity to resist.

The qualitative discontinuity between *tranq sleep* and a typical opioid nod was articulated clearly by all participants who spoke to the topic. “Totally different than a nod. It’s sleeping. You’re going to sleep, you’re tired,” bemoaned one participant (White, Female); while another offered an equally terse comparison: “Not like a nod. A nod you’ll gradually go into, okay, but the xylazine, it just puts you out. Like you don’t realize that you’re going to sleep, and then you just wake up” (Asian, Female). Several participants described the experience as akin to an alcohol-induced blackout. One participant awoke to discover her clothes folded with no memory of doing so, underscoring the blackout-like memory loss associated with *tranq sleep*.

*Tranq sleep* onset was described as sudden and profound. Participants characterized the transition from consciousness to unconsciousness as acute and inescapable. “Almost instantly, when you smoke it [xylazine] and blow out, you’re nodding [*sic*] and you can’t fight that nod—it don’t matter where you’re at” (White, Male). Several participants reported how the strategy for counteracting fentanyl sedation with methamphetamine was ineffective at combating *tranq sleep*, further distinguishing it from the fentanyl effect.

Participants demonstrated wide variability in the duration of a ‘typical’ *tranq sleep* event, with reports ranging from five minutes to three or more hours. Similar to alcohol-induced blackouts (Merrill et al., 2022), many noted having no reliable way for determining how long they had been unconscious. While this unpredictability was itself a source of concern, the inability to function during *tranq sleep* was among the most commonly cited reasons for disliking xylazine. As one participant woefully explained: “I really don’t care for it. I feel like I’m just wasting days away, you know what I’m saying; like I don’t do anything productive. At least whenever I was doing the other stuff [heroin] I still would get up and clean my house or do my laundry or something. Now I can’t even move from the bed” (White, Female). For participants who initiated opioid use with semi-synthetic opioids like oxycodone and/or heroin, the contrast with *tranq sleep* was particularly striking. Heroin was consistently recalled not only as compatible with the ordinary requirements of daily functioning—going to work, managing household tasks, caring for children—but as actively motivating participants to engage in those activities, lending energy and purpose to daily life in ways that made routine demands feel manageable. Fentanyl, by contrast, was disparaged for rendering this same routine activity extremely challenging, even in the absence of xylazine. Xylazine’s presence, however, serves to compound this sedative burden further, eroding participants’ motivation, capacity to function, and locus of control in ways that heroin, even at peak physical dependence, had not.

Safety concerns are thus an inseparable dimension of *tranq sleep*. Unhoused participants or those with housing instability described feeling acutely vulnerable during *tranq sleep* episodes, becoming unable to either perceive or respond to threats in their environment. “I don’t want to be tranquilized,” lamented a woman in her forties. “I don’t want to be not able to move or function, especially being homeless, you know what I mean. I don’t want to be in my tent and not be able [to act] if someone’s coming up [on me]. I need to be aware of what’s going on around me all the time” (White, Female). Another participant spoke to the fear of tranq sleep while being unhoused and isolated in a tent: “They can come in and rob you or whatever and you’re not hearing nothing. Or if you do hear it you’re not doing nothing, you can’t do nothing really” (White, Male). Additional safety concerns surfaced in the context of employment and driving. Participants aggrieved the extent to which *tranq sleep’s* unpredictability precluded them from social functioning, holding down a job, or operating vehicles.

The most clinically significant feature of *tranq sleep* was the acute onset of opioid withdrawal upon awakening. Many participants described waking from *tranq sleep* feeling severely sick, as though no pharmacologically meaningful time had elapsed despite having been sedated for an extended period. “Yeah, because that stuff is so tranquilizing and when you wake up you’re sick immediately. It’s almost like you never had any fentanyl. You were sick but you couldn’t tell because you were asleep” (White, Female). Another participant described this experience in plain terms: “You wake up in full withdrawal and, holy shit, now I got to get some more. People hate that, that’s why people don’t like tranq either— because you sleep through what should have been your time to be well and you come back [to consciousness] and you’re sick” (White, Female).

Several participants characterized *tranq sleep* withdrawal as markedly worse than fentanyl withdrawal by itself. “The tranq is, it’s different, it’s horrible, it’s worse, it’s worse than the fentanyl sickness. The tranq just lays me down. It makes it to where I just want to sleep and do nothing but sleep. And when you wake up you feel ten times worse than what you would feel if you woke up off fentanyl” (Asian, Female). Several participants reached for emphatic language to convey the severity, describing waking up “already dope sick” or waking up “in the worst of it.” This pattern was offered by participants as a recurring explanation for why xylazine adulterated fentanyl was distinctly worse than unadulterated fentanyl for maintaining social functioning, and why nearly everyone maintained a clear animus towards xylazine

### Discernment

Participants described several sensory strategies for identifying xylazine in fentanyl, with taste, smell, and the physical behavior of the drug when smoked discussed most frequently and consistently across interviewees.

The predominant discernment strategy was taste and smell when burned, with more than a third of interviewees identifying taste as their primary method for detecting xylazine. While fentanyl was described as having a sweet or medicinal taste when smoked, participants most frequently characterized xylazine-adulterated fentanyl as producing a strong, unpleasant, and distinctively chemical taste. Xylazine was repeatedly described as “bitter,” “harsh,” “nasty,” and tasting like “a mouthful of chemicals” when burned.

One participant offered a more nuanced account, describing xylazine’s taste as paradoxically sweet but unpleasant: “It’s got a sweetish, nasty taste. It’s a funny looking color. It’s almost, to me, when you first heat it up, you first taste it, it’s like you’re heating up powdered or granulated sugar. It’s got a sweet taste to it” (White, Male). This refined illustration stands in contrast to the more common “bitter” or “chemical” characterization and may reflect variation in formulation or concentration across different arms of illicit supply chains, or even a diversity of palate sensitivity with taste as a sensory strategy.

Although most participants insist there is no reliable or consistent way to visually detect xylazine’s presence in fentanyl, the physical presentation and textural consistency of fentanyl products were frequently discussed as indicators of purity and adulteration more broadly. Fentanyl consumers expressed a preference for “chunk” over powder, with loose, powdery consistency interpreted as evidence it was “stepped on.” “I like smoking chunk because there’s a better chance that my shit’s not cut. As to where if it’s powder, they’re stepping on it; they’re cutting it” (White, Female).

The pyrolytic behavior of fentanyl when heated on tinfoil was one of the most consistently reported discernment strategies. Most participants who smoked fentanyl insisted on knowing when it was unadulterated (i.e., without xylazine) by the way it eventually turns into a yellowish color and by how smoothly it runs across the foil surface: “If it’s real good fentanyl it’ll turn like a goldish champagne color and it’ll be real clear. When the white, powdery chunk or the white powder starts melting, it’ll have like a pinkish-purple tint to it, it’ll go clear and then like a champagne color” (Male, White).

In sharp contrast, most descriptions of *xylazine*-adulterated fentanyl depict it as turning into a bubbling liquid that volatilizes instantly, leaving behind a black, burnt residue that cannot be again vaporized. Unlike unadulterated fentanyl that liquefies into a stable, viscous oil, *tranq fentanyl* was described as surprisingly unstable and wholly incapable of “running” back-and-forth across the foil. “When you smoke it [xylazine] you can taste the taste of it, but also, like when you’re burning it, it’s [xylazine’s] going to burn a little bit black, or darker black—it bubbles up” (Female, White).

Participant accounts varied regarding whether xylazine burned at a different rate from fentanyl, with some reporting flash combustion and others rejecting this observation. In a notable behavioral adaptation, multiple participants reported adding methamphetamine to a “foil pack” of fentanyl to optimize its viscosity so it ‘runs’ better on foil. This strategy serves to both improve the overall smoking experience and to counteract xylazine’s tendency to burn unfavorably on foil. As one participant tells: “I always mix my meth in it [xylazine-adulterated fentanyl] so it runs more of a honey color anyways, but it leaves like a resin on it [the foil]” (Female, White).

Several participants described performing a deliberate “test smoke” to ascertain whether their fentanyl was adulterated with xylazine before consuming a preferred amount. This emic practice involves heating a small portion of fentanyl on foil to observes how it burns, tastes, smells, or “hits” [vaporizes] before committing to a full dose. “To me the best way to test it without a [xylazine] testing kit [strip] is to put it on some foil and just melt it real quick” (Male, White). One participant who preferred injecting fentanyl to smoking portrayed “test smoking” as a rapid screening strategy for safer injecting, even though smoking was not his preferred route of fentanyl consumption. “Even though I’m a shooter, I do still smoke. But if I don’t have time to do a shot and try it out, the best way to test it is to throw the piece [of fentanyl] on the foil [and] make a run with it. And depending on how it burns, if it leaves black shit, or trash, you know it’s no good” (Male, White).

A meaningful minority of participants reported being unable to detect xylazine’s presence in fentanyl by sight, most commonly in relation to a product’s visual appearance, but also taste, with some participants stipulating that xylazine has no distinct taste or smell when smoked. “I don’t think it has a taste, really. I think it’s more of a reaction, has more of a reaction. The fentanyl has a taste but that [xylazine] doesn’t really give a taste, I think it just helps with the feeling” (Male, Black). Others noted that the only reliable signal is the physical effect the drug produces when ingested. “The only difference is, you just wake up [from *tranq sleep*] and know that you just hit it [xylazine]” (Female, White).

Among participants who preferred injecting fentanyl, some described how the fentanyl solutions they fashioned became thick and viscous and difficult to draw into a syringe, a type of hydrogelation reminiscent of prescription opioids (e.g., Opana) they attributed to xylazine. One participant described how her fentanyl, which was previously clear and homogeneous in aqueous solution, now turned white, cloudy, and very thick in the cooker. “Since the xylazine came in I’ve been noticing that when I put flame to it and then take the flame away, the white comes back; it turns white again or it gets really thick again” (Female, White).

Across sensory strategies, xylazine’s physical and pharmacologic effects emerged as the most reliable indicators of its presence. Participants identified acute onset of unconscious sedation as indicative of xylazine exposure, with some reporting sedation so instantaneous they only knew it was xylazine upon awaking. “You hit that shit, when you start pushing [the plunger and injecting] it, you’ll be sitting there looking and then the next thing you know you’re either waking up because they hit you with Narcan [naloxone] and you’re lucky to come back, or you wake up dead. That’s pretty much all it is” (Male, White).

### Likes and Dislikes

We asked participants what they liked and disliked about xylazine. Nearly all participants strongly disliked xylazine, with only a few accepting it as a welcome addition to a fentanyl supply that no longer gets them high. Many dislikes were directly tied to *tranq sleep*, such as the inability to function, concerns about personal safety, and awakening in acute withdrawal. A frequently mentioned aversion to xylazine was its unfavorable contrast to a typical opioid high. Participants described the opioid (read: heroin) high as a “warm bath,” or a euphoric and energizing experience, and something they needed to function better at work or with family; in contrast to xylazine that was characterized as less rewarding, less energizing, and more sedating. “I always liked heroin. Even when I had to do a lot of it, I still liked it. It was like slipping into a warm bath. This [xylazine-adulterated fentanyl] is cold and clinical. Great, I’m well, but oh my god” (Female, White).

Other frequently cited dislikes of xylazine include awakening from *tranq sleep* in peak withdrawal, fear of naloxone not reversing a fentanyl overdose, and the risk of acquiring tranq wounds. Participants who attributed wounds to xylazine specified they were more numerous with atypical characteristics compared with abscesses they had experienced in the past. “I think people are still scared of the tranq, and scared because they know. I mean, we have wounds all over us and it’s obvious that it’s not just fentanyl” (Female, White).

Despite the preponderance of negative responses, it is notable that a select few of participants expressed positive regard for xylazine. This handful of consumers enjoyed the way tranq amplified the fentanyl high that their tolerance had made harder to attain. “Yeah, [the xylazine is] a positive attribute because the fent ain’t doing nothing for me anymore, it’s just keeping me well where I’m not sick as a dog. As the xylazine with the fent, I know I won’t be sick if it has fentanyl in it. But with [xylazine] I will actually get the enjoyment of doing it [fentanyl], the euphoria of it [the combination]” (Female, White). Some participants described the desire for xylazine vicariously, noting that others in their networks actively sought xylazine adulterated fentanyl. “My cousin told me he wished he could find just the xylazine by itself. And I was like, ’Why?’ And he’s like, “because I love it and miss it. This [fentanyl] dope here just ain’t doing it for me no more’” (Male, White).

## DISCUSSION

The purpose of this qualitative study was to examine how fentanyl consumers in west-central North Carolina detected and responded to xylazine’s presence in the fentanyl supply. Consistent with qualitative research conducted in Northeast and Mid-Atlantic states, participants expressed strong and near-universal aversion to xylazine (Reed et al., 2022; Reed et al., 2025a, 2025b; Spadaro et al., 2023; Eger et al., 2024). The current study advances the existing xylazine literature by documenting a sophisticated repertoire of pre-consumption sensory discernment strategies—test smokes, foil burn behavior, taste, smell, and viscosity assessment—that participants actively employed to detect xylazine. Prior qualitative work has captured post-hoc recognition of xylazine through its post-consumption effects (Reed et al., 2022; Eger et al., 2024), but preemptive, pre-consumption detection strategies of this kind have not previously been documented in the literature. By documenting these strategies in a rural fentanyl market, this study adds both geographic scope and phenomenological depth to a nascent body of public health literature on sensory strategies for discerning street drugs.

The most notable finding to emerge is a phenomenon we refer to as *tranq* sleep. Tranq sleep is a protracted state of profound sedation that is rapidly induced after xylazine is smoked or injected, lasting anywhere between several minutes to several hours. Participants reliably distinguished *tranq sleep* from both the familiar heroin nod and heavy fentanyl sedation. Understanding what makes *tranq sleep* phenomenologically distinct from these prior reference points requires briefly reconstructing both.

Participants consistently described losing consciousness immediately after consuming xylazine (via smoking or injecting) without any early warning signs and awaking in positions that made clear they had been caught off guard and unconscious for much too long to efficiently resume pre-sleep activity. Others recounted awakening in public or semi-public settings with no memory of how they got there or how much time had passed, describing the experience as a kind of forced disappearance from their own lives. Several participants reported waking to find that they had been robbed while *tranq sleeping*, or unable to account for valuables—or in some cases their own bodies. *Tranq sleep*’s unarousable and protracted character was the distinguishing feature participants returned to most consistently when discussing xylazine’s distinguishing characteristics.

Another distinguishing feature of *tranq sleep* is the experience of awakening in advanced-stage fentanyl withdrawal. Participants were virtually unanimous in their depictions of what it felt like to come out of tranq sleep and all drew explicit contrasts with the way they had come to understand recovery from other opioid states. Emerging from the euphoric serenity of a heroin ‘nod’ carries with it a continuity, participants explained, that persists into wakefulness, as the person nodding resurfaces into a body that still feels adequately medicated and capable of re-orienting and resuming the ordinary activities of their day. Fentanyl sedation occupied an intermediate position as an involuntary descent into brief unconsciousness followed by a period of disorientation that lifted quickly, with enough residual drug effect to forestall onset withdrawal, allowing consumers to reorient, however unsteadily, to their surroundings. Compared to *tranq sleep*, fentanyl’s deep sedation was still recognized as manageable because of how quickly consumers return from episodic fentanyl inertia to a mostly functional state. Current study participants knew both opioid states intimately well and were unambiguous that *tranq sleep* was neither.

Participants described wakening from *tranq sleep* not with lingering euphoria (heroin) or the capacity to reorient (fentanyl) but into the full and unambiguous misery of late-stage withdrawal: muscle cramps, bone-deep aching, profuse sweating, nausea, and a physical agitation that participants consistently distinguished from the early warning signs of ordinary fentanyl withdrawal. Current study findings suggest that xylazine imposes an interlude of forced detoxification during which fentanyl metabolism continues unabated while the consumer remains insensate and unable to re-dose. This appears to defer early withdrawal episodes so they continue unimpeded in the unconscious (physically dependent) consumer until they awake and experience the pangs of compounded and postponed fentanyl withdrawal.

The withdrawal participants described upon awakening from *tranq sleep* was characterized in terms consistent with late-stage fentanyl withdrawal: immediate, acute, and severe enough to compel re-dosing without delay. Whether awakening withdrawal (i.e., tranq sleep withdrawal) represents a compounded form of fentanyl-only withdrawal, or a more pharmacologically complex, synergistic withdrawal involving co-occurring physical dependence (both fentanyl and xylazine) is a research question this study is not equipped to answer. Repeated and involuntary xylazine exposure from a xylazine-saturated fentanyl supply creates the pharmacological conditions under which physical dependence on xylazine could develop in parallel with fentanyl dependence, with withdrawal from both substances interacting in ways that can intensify and accelerate the severity of *tranq sleep withdrawal*. Participants themselves, however, did not describe what they experienced as a novel or unfamiliar type of withdrawal. They described it as fentanyl withdrawal, but the possibility that xylazine withdrawal was operating simultaneously and synergistically, amplifying the fentanyl withdrawal signal into something more severe and more immediate than fentanyl withdrawal alone would produce, cannot be ruled out. The neurobiological interaction between alpha-2 adrenergic agonist withdrawal and synthetic opioid withdrawal is poorly understood in the context of concurrent street-level exposure. Current study participants described with such consistency the harrowing nature of *tranq sleep withdrawal* that it remains an important and unanswered clinical and pharmacological question.

The immediate, compulsory nature of re-dosing upon awakening from *tranq sleep withdrawal* was described by participants in terms less volitional than physiological: participants did not wake into respite but into active, late-stage withdrawal already severe enough to demand immediate remedy. The mechanistic logic that emerges from these accounts is a cycle as self-reinforcing as it is inescapable: tranq sleep imposes an interval of unconsciousness during which fentanyl withdrawal advances uninterrupted, and the only available remedy upon waking—in a xylazine-saturated supply—is the drug that begins the cycle again. Participants did not use the language of pharmacological cycling to describe this process, but the mechanism they articulated with consistency and specificity was unmistakable. The coherence with which participants across this study described this pattern positions *tranq sleep* not as an incidental side effect of xylazine exposure but as a proximate and self-perpetuating mechanism of harm, with direct implications for daily consumption frequency, cumulative drug exposure, and swelling toxicity. This dynamic compounds the already well-documented problem of fentanyl’s short duration driving higher injection frequency (Duhart Clarke et al., 2022; Lambdin et al., 2019; Mayer et al., 2018) and adds a second, xylazine-specific, cycling mechanism operating in parallel. This finding is consistent with prior qualitative work documenting worsened withdrawal as a distinctive feature of xylazine exposure (Spadaro et al., 2023; Reed et al., 2025a; Eger et al., 2024).

The functional consequences of *tranq sleep* that followed from this cycle—the inability to work, care for children, maintain housing, or respond to physical threats while incapacitated—emerged in this study as the dominant driver of participants’ aversion to xylazine. Taken together, convergent findings across Philadelphia, Miami and now a rural region in North Carolina constitute an emerging and geographically broadening epidemiologic picture of *tranq sleep* as an adverse consequence of xylazine exposure. A public health response will require a holistic approach including clinical protocols that account for *tranq sleep’s* severe withdrawal trajectory, harm reduction training calibrated to the lack of warning before losing consciousness, and structural interventions capable of reducing the vulnerability of consumers (e.g., unhoused persons, unaccompanied women) who experience *tranq sleep* in unsheltered or otherwise hidden environments (e.g., wooded encampments, squats).

Beyond using *tranq sleep* to indicate xylazine exposure, participants demonstrated a sophisticated knowledge of their drug supply and employed a number of sensory strategies (e.g., taste, smell, visual inspection) to detect xylazine in fentanyl. In a manner consistent with the homegrown drug discernment practices previously documented among PWID in this region (Duhart Clarke, 2022), participants in the current study functioned similarly as lay chemists, systematically dissecting sensory information from each consumption event to assess the composition of an increasingly unpredictable supply. Prior research from North Carolina established how illicit opioid consumers regularly employ sensory strategies to identify sudden changes in the drug supply using physical appearance and physiological effects as interpretive signals, particularly in the absence of any formal means of verification (Zibbell et al., 2021). The sensory strategies that helped heroin consumers detect fentanyl in the early fentanyl era are now being applied by fentanyl consumers to detect xylazine in the late fentanyl era, extending a well-documented capacity for informal drug discernment to the latest adulterant to enter the supply.

Consumers across multiple studies have described applying similar strategies to detect xylazine, using onset, potency, duration, and its distinctive sedation profile as cues to adulterant presence (Reed et al., 2025b; Park et al., 2024; Reed et al., 2022). In the current study, several participants described combining sensory cues into a deliberate *pre-consumption* screening practice referred to as a ‘test smoke.’ Performing a test smoke is analogous to the much revered “test(er) shot” and represents an analogous harm reduction strategy designed by PWID who became fentanyl smokers. Because xylazine’s presence in regional drug markets is rarely disclosed by street-based sellers, these sensory discernment strategies function as a form of informal, sentinel surveillance that no official drug surveillance system has yet to match with comparable speed or granularity.

Consistent with the broader qualitative literature, virtually all fentanyl consumers in the current study expressed a general animus toward xylazine, with the terms of dislike strikingly consistent across participants. Participants repeatedly invoked the warm, enveloping quality of a typical opioid high as the standard against which xylazine (and fentanyl) sedation was found profoundly wanting. Xylazine was described as cold and clinical and far more akin to fentanyl’s deep sedation than the energetic reward commonly associated with heroin and other semi-synthetic opioids (e.g., oxycodone, hydromorphone). The contrast between opioid-induced energy and sedative lethargy is not merely metaphorical but serves to illuminate the pharmacological basis for opioid consumer’s aversion to sedatives as a substitute for or adulterant of full μ-receptor agonist opioids. Here, the distinction drawn by participants was not between sedation or no sedation but between two qualitatively different depressant states: one experienced as euphoric, energizing, and motivating and the other as blunt, incapacitating, and empty.

The primary drivers of this near-universal aversion to xylazine that emerged from participants’ accounts above—including functional incapacitation from tranq sleep; safety vulnerabilities during bouts of unconsciousness; an inferior high relative to heroin; an accelerated and compressed withdrawal burden upon awakening and a pervasive fear of overdose—are consistent with findings across multiple qualitative studies in a diversity of settings. Reed et al. (2022) found that none of their Philadelphia participants wanted xylazine in their supply and its sedating effects were explicitly linked to fears of victimization and theft during unconsciousness. Reed et al. (2025a) subsequently documented withdrawal burden and safety vulnerabilities as the foremost concerns among Philadelphians in focus group discussions. Correspondingly, Eger et al. (2024) reported from Miami that SSP participants routinely depict xylazine as an unwanted contaminant that disrupts daily functioning and psychosocial well-being. Altogether, the current study contributes a rural, southern context to this convergent body of urban evidence which the same constellation of aversions emerged with comparable specificity. This convergence strongly suggests that the harms participants describe are not artifacts of urban market conditions but features of xylazine exposure itself generalized across geography, consumption modes, and illicit opioid markets.

Yet, even with the virtually unanimous hostility towards xylazine among current study participants, the positive regard expressed by a nominal yet meaningful minority deserves careful analytic attention rather than dismissal. This finding is consistent with other US studies (Spadaro et al., 2023; Friedman et al 2022) indicating a minor heterogeneity in consumer preference for xylazine. These participants valued xylazine primarily for its capacity to amplify a fentanyl high that tolerance and market volatility had rendered inadequate. In this context, xylazine provides the ability to restore, however temporarily and incompletely, some approximation of ‘the opioid effect’ that fentanyl alone could no longer produce for this outlier group. Combined with current findings, this growing body of literature suggests that xylazine preference is shaped, in part, by fentanyl’s own pharmacologic limitations, with its exceedingly short duration creating demand for adulterants that can extend the window of fentanyl intoxication, even at the cost of adverse consequences for whom the vast majority of fentanyl consumers find intolerable.

This study has several limitations. The sample was small (N = 25) and purposively recruited through a community-based, syringe services program (SSP) in west-central NC, so findings are not generalizable to the broader population of people who use drugs in North Carolina or elsewhere. Study participants were predominantly non-Hispanic White and reflected the demographic profiles of SSP participants in this region of NC, but such uniformity may limit generalizability to communities of color impacted by xylazine in other regions. Recall bias is inherent in retrospective accounts of drug use experiences, but this was partially mitigated by eligibility criteria requiring recent drug use (past 48 hours) and by an in-depth interview format that allowed participants to clarify accounts across the sequence of extended conversation.

Because the interview guide was developed prior to the full emergence and naming of certain xylazine-related phenomena (e.g., tranq sleep), some dimensions of participants’ experience may not have been as comprehensively explored as they would be in an instrument developed with foreknowledge of this phenomenon. Still, this constraint is offset, in part, by years of immersive fieldwork in this region of rural NC, with prolonged research with impacted communities establishing a deep level of trust and rapport that is critical for tracking emerging drugs and nascent harm. Accordingly, a survey investigation was subsequently conducted in this region of NC so future analyses with quantitative instruments can capture and confirm these qualitative findings.

### Public Health Implications

Findings of this qualitative study carry urgent implications for harm reduction and public health. *Tranq sleep* and ‘waking withdrawal syndrome’ represent discrete and underrecognized harms that existing overdose prevention frameworks were not designed to address. Naloxone distribution and conventional overdose response training remain essential but neither is calibrated to the specific demands *tranq sleep* places on bystanders, peers, and harm reduction workers. The people most likely to be present during a *tranq sleep* episode cannot respond effectively to a phenomenon they have not been told to expect. Closing this information gap is a first obligation of public health. Equally important is what our findings reveal about the capacity of street drug consumers to function as active contributors in sentinel surveillance efforts. The sensory discernment strategies documented here represent contributions of particular significance. ‘Test smoking’ is a harm reduction practice identified here for the first time in the published literature. In effect, test smoking is a consumer-generated drug checking tactic developed in the absence of formal testing infrastructure (Alexander et al., 2022) and deserves recognition as such. Formalizing discernment practices and training consumers to systematically document market changes would extend the reach and speed of actionable surveillance in ways that no laboratory or forensic toxicology dashboard can match (Zibbell et al., 2023). As this study and its predecessors demonstrate, people who consume street drugs can identify adulterants, characterize pharmacologic effects with lived precision, and communicate actionable intelligence on emerging threats before they appear in overdose mortality data. The cost of not formalizing consumer surveillance capacity is measured in the number of unwitting people who get sick or die from toxic exposures that seasoned consumers already knew were there.

**Table 1.** Participant Exemplar Quotes by Theme.

| Theme | Exemplar Quote |
| --- | --- |
| Tranq Sleep | "Big nod, bro, and that's what people, you know, they see you nod and they think you're high but really it's just putting you to sleep." (Male, White, Gaston County) |
| Tranq Sleep | "It's not really even a high, it's you, just you're out, you go to sleep." (Female, White, Gaston County) |
| Tranq Sleep | "But you can like, you can shake it to an extent, like if somebody comes in or whatever and you hear them, you're going to lift your head up at least. The xylazine, it'll be like, they can come in and rob you or whatever and you're not hearing nothing. Or if you do hear it you're not doing nothing, you can't do nothing really." (Male, White, Gaston County) |
| Tranq Sleep — Hits Instantly | "Almost instantly when you smoke it and blow out you're nodding and you can't fight that nod, it don't matter where you're at." (Male, White, Gaston County) |
| Tranq Sleep — Inability to Function | "It knocks you out and you can't hardly get off of it, it just makes you like a damn mummy and, because you can't, really can't function off the shit. You can't, it ain't like, heroin you can function off, you can go to work." (Male, White, Gaston County) |
| Tranq Sleep — Waking Up in Withdrawal | "Through your withdrawal and you wake up in full withdrawal and, holy shit, now I got to get some more. You know, and people hate that, that's why people don't like tranq either, because you sleep through what should have been your time to be well and you come back and you're sick." (Female, White, Gaston County) |
| Tranq Sleep — Waking Up in Withdrawal | "Now I wake up and I'm, no matter what, if you fuck around and nod and doze off and you wake up, you just, you're going to wish you had none because automatically, you're going to be sick automatic, it's like you wake up from a tranquil state, boom, and you're sick automatic." (Male, Black, Gaston County) |
| Xylazine Withdrawal is Worse | "The tranq is, it's different, it's horrible, it's worse, it's worse than the fentanyl sickness... the tranq just lays me down, it makes it to where I just want to sleep and do nothing but sleep. And when you wake up you feel ten times worse than what you would feel if you woke up off fentanyl." (Female, Asian/White, Burke County) |
| Xylazine withdrawal is worse, with burning of skin | "I noticed a big difference in, in the skin withdrawals particular, and the burning, like it feels like your skin's burning...It feels like if you had, like, one of those space blankets and it sticks to your skin; imagine it sticking to your skin hot." (Female, White, Burke. Co) |
| More Likely to Overdose with Xylazine | "You hit that shit, when you start pushing it, you'll be sitting there looking and then the next thing you know you're either waking up because they hit you with Narcan and you're lucky to come back or you wake up dead." (Male, White, Gaston County) |
| Taste of Xylazine Smoke | "From my experience with it, it burns like black or like a darker color." (Female, White, Burke County) |
| Taste of Xylazine Smoke | "It's harsh and it stunk and it tasted like shit." (Male, White, Gaston County) |
| Taste of Xylazine Smoke | "You don't want to taste sweet, you know that's cut with sugar. Yeah, if you taste sweet you've got trash." (Male, White, Gaston County) |
| Taste of Xylazine Smoke | Int: And what was the biggest sign that you knew it was xylazine or something else?<br>Participant: "Taste. The taste, it had an odd taste to it and it has an odd odor to it. And once you hit it the taste, you can just instantly taste. But more than anything you're going to like instantly nod with a xylazine." (Female, White, Gaston County) |
| Appearance of Fentanyl and Xylazine When Smoked | "A good fent will be like a yellowish or whatever, I mean, after a little while — it'll be like a real yellowish, maybe a tan color, light brown if anything. But no, xylazine, it's like it burns, I don't know, it turns black." (Male, White, Gaston County) |
| Appearance of Xylazine After Burning | "After you burn it and it starts going to where it's black like at the end, it starts turning black and stuff like that, that's where you know if it's cut." (Male, White, Gaston County) |
| Drugs Thickening When Heated Then Cooled | "It seems more a little like thick than just water but it's clean, it doesn't look like anything's in it, it doesn't like crystallize up in the syringe, but it just seems like a thicker consistency to push through." (Female, White, Gaston County) |
| Negative Discernment | "I don't think it has a taste really, I think it's, it's more of a reaction to it, it has more of a reaction, I don't think it has a taste. The fentanyl has a taste but that doesn't have a, I don't think it really gives a taste, I think it's just the feel, that it helps with the feeling." (Male, Black, Gaston County) |
| Suspected Xylazine Overdose | "I OD'd and died two weeks ago and it was because of the stuff that this guy had... as soon as I smoked it, it numbed part of my head and then it made me very hot and I broke out into a sweat, and fentanyl doesn't do that... And as soon as I hit it, it was just like a big whoosh. And so I stood up and I went and walked over to the couch and I sat down and I fell out and I died." (Female, White, Gaston County) |
| Likes Xylazine — Stronger High | "My cousin told me, he said he wished he could find just the xylazine by itself... he loves it and he misses it, he's like, "This dope here just ain't doing it for me no more." (Male, White, Gaston County) |
| Likes Xylazine — Stronger High | "Yeah, it's a positive attribute because the fent ain't doing nothing for me anymore, it's just keeping me well where I'm not sick as a dog, as where the xylazine with the fent, I know I won't be sick if it has xylazine in it. But I will actually get the enjoyment of doing it, the euphoria of it." (Female, White, Gaston County) |
| Likes Xylazine — "Clear" Rush | "It'll make it feel like to me, you know, like a clear rush, you know what I mean... it's like a relaxing heat to your body." (Male, White, Gaston County) |
| Likes Xylazine — Energy | "Yeah, it gets you going, yeah, it gets you going. But you're actually, you know what I'm saying, you're more relaxed, you're more up and alert." (Male, White, Gaston County) |

## Data Availability

Some data may be available upon reasonable request of the authors.

